# Multiethnic radiogenomics reveals low-abundancy microRNA signature in plasma-derived extracellular vesicles for early diagnosis and molecular subtyping of pancreatic cancer

**DOI:** 10.1101/2024.11.22.24317764

**Authors:** Wenjie Shi, Jianying Xu, Yi Zhu, Chao Zhang, Julia Nagelschmitz, Maximilian Doelling, Sara Al-Madhi, Ujjwal Mukund Mahajan, Maciej Pech, Georg Rose, Roland Siegfried Croner, Guo Liang Zheng, Christoph Kahlert, Ulf Dietrich Kahlert

**Affiliations:** Molecular and Experimental Surgery, Clinic for General-, Visceral -, Vascular- and Transplantation Surgery, Medical Faculty and University Hospital Magdeburg, Otto-von-Guericke University, Magdeburg, Germany; Department of Medicine II, Hospital of the LMU Munich, 81377, Munich, Germany; Department of Gastroenterological Surgery, The Affiliated Hospital of Jiaxing University, Jiaxing, Zhejiang, China; Department of Radiology, The First Affiliated Hospital of Wannan Medical College, Wuhu City, Anhui Province, China; Clinic for Radiology and Nuclear Medicine, University Hospital Magdeburg, Magdeburg, Germany; Research Campus Stimulate, Otto von Guericke University Magdeburg, Germany; Department of Gastric Surgery, Cancer Hospital of China Medical University (Liaoning Cancer Hospital and Institute), Shenyang, China; Department of General, Visceral and Transplantation Surgery, Heidelberg University Hospital, Heidelberg, Germany

**Keywords:** PDAC, EV, microRNA, CT, radiogenomics

## Abstract

**Purpose:** Pancreatic cancer is a highly aggressive tumor, and early diagnosis and treatment significantly improves the clinical benefit for patients. However, currently there is a lack of effective methods to accurately detect this disease. In our study, we develop a liquid biopsy signature of EV miRNAs based on associated radiomics features of patients’ tumors in order to provide new insights for the early diagnosis of pancreatic cancer.

**Experimental Design:** A total of eight datasets enrolled in this study, featuring clinical and imaging data from different benign pancreatic lesions and malignant pancreatic cancers as well as small RNAseq data from cargo of plasma extracellular vesicles of tumor patients. Radiomics Feature Extraction and different features analysis performed by limma packages. Feature selection was performed by Boruta algorithms and radiomics related signature model was build and validated by lasso regression algorithms. Radiomic signature related to low abundance EV miRNAs was analyzed by weighted gene co-expression network analysis. The diagnosis ability of above miRNA are validated by ten machine-learning algorithms. The shared target of candidate miRNAs were predicted and clustered followed by subsequently probing for predicting survival benefit of the patient, drug sensitivity of tumor cells and functional differences.

**Results:** A total of 88 significant radiologic features demonstrate differences between benign lesion and pancreatic cancer. Three radiomics factor related signature related a plasma EV-miRNAs triplet possessing high accuracy in diagnosis cancer from benign lesions. Moreover, clustering miRNA and there predicted molecular signaling partners in tumor tissue identified tow molecular subtypes of pancreatic cancer. Cluster stratification separates low risk tumors in terms of severely prolonged overall survival time of patients, higher sensitivity to immune therapies. We also propose the potential of purposing selected targeted drugs to specifically targeting the molecular activation markers in high-risk tumor cluster.

**Conclusion:** Our three radiogenomics related blood plasma extracellular vesicle microRNA signature is a useful liquid biopsy tool for early diagnosis and molecular subtyping of pancreatic cancer, which might treatment decision making.

**Statement of translational relevance:** The identification of a low-abundance microRNA signature in plasma-derived extracellular vesicles offers significant translational potential for the early diagnosis and subtyping of pancreatic cancer, particularly across diverse ethnic populations. This discovery could lead to the development of non-invasive liquid biopsies that improve early detection rates, a critical need for a cancer with notoriously poor prognosis due to late diagnosis. By incorporating this microRNA signature into clinical practice, oncologists may be able to detect pancreatic cancer at earlier, more treatable stages, enhancing patient survival rates. Additionally, the subtyping capability of this signature could guide personalized treatment strategies, allowing for more targeted therapies based on specific cancer subtypes. This could ultimately reduce the need for invasive diagnostic procedures and optimize treatment efficacy, reducing adverse effects and improving outcomes. The integration of radiogenomics and liquid biopsy technologies promises to be a powerful tool in the future of cancer medicine, particularly in underserved populations.

## Introduction

Pancreatic cancer is one of the most lethal tumors having a poor prognosis and patients suffering from this disease show one of the lowest 5-year overall survival rate of all cancer patients with approximately 13%. One of the main reasons for this dismissal prognosis is the lack of a proper early detection possibility, resulting in late diagnosing often in advanced, metastatic stage^1^

The detection and diagnosis of pancreatic cancer, of which approximately 90% are classified as pancreatic ductal adenocarcinoma (PDAC), currently relies primarily on modalities of medical imaging, such as computer tomography, magnet resonance imaging, positron emission tomography and transabdominal ultrasonography^2^. The most common biomarker considered for PDAC differential diagnosis is elevated blood abundancy of carbohydrate antigen 19-9 (CA19-9) and carcinoembryonic antigen (CEA), though these are only used as prognostic markers and not effective for screening or early diagnosis^3^ Nowadays, new markers based on liquid biopsy such as microRNA are discerned and might pose as promising tools for early detection of PDAC. miRNA are non-coding RNAs that target genes and regulate their expression by inhibiting mRNA translation or enhancing their degradation^4^. Currently, extracellular vesicles (EV) are gaining attention as disease specific marker since they carry the material of their secreting cells and are therefore considered to contain tumor-derived elements, showcasing their molecular fingerprint (Bamankar und Londhe 2023). It has been shown multiple times that miRNA derived from small EVs play a role in differentiation and metastasis of cancer^5,6^. The rapidly developing field of data mining and analytical techniques provide new insights and make the discovery of relevant key players more feasible. Numerous miRNA have been described using co-expression network analysis that might be applicated as diagnostic or prognostic biomarker, for patient stratification or disease recurrence^7^.

Benefiting from interdisciplinary advances in artificial intelligence, the integration of machine learning and genomics has led to breakthroughs in the early diagnosis and classification of tumors. For example, we used machine learning algorithms to assist in the development of a three-serum miRNA signature that effectively provides early warning of premalignant pancreatic cancer^8^. Another model, based on machine learning algorithms, focuses on the immune subtypes of triple-negative breast cancer, offering critical insights for identifying patients who may benefit from immunotherapy^9^. In the current study, we employed radiogenomics technology, another product of interdisciplinary collaboration between medicine and engineering. This novel approach integrates the quantification of image features from CT or MRI, which are then correlated with genomic signatures and allows for a non-invasive prediction of molecular characteristics^10^. Such as, claim to be able to predict the occurrence of p53 mutations using CT images by radiogenomic analysis and hence make a statement on the prognosis^11^.

In the present study, we aim to develop a liquid biopsy signature of EV-derived miRNA based on the radiogenomic analysis of CT images derived from ethical diverse background and interrogating small RNAseq data of plasma-derived total EVs, in order to advance the diagnostic possibilities and the molecular subtyping of PDAC.

## Materials and Methods

### Data resources and pre analytics preparation

A total of four hospitals and four public datasets, comprising a total of eight datasets enrolled in this study, University hospital Magdeburg in Germany (UMMD), and Jiaxing Hospital Center, China (JHC), provided enhanced computed tomography (CT) images of 46 intraductal papillary mucinous neoplasm (IPMN) patients and 127 pancreatic cancer (PC) patients as training dataset. For test datasets, CT data resource from Wanan medical university hospital (WUH) with 27 PB patients and 72 PC patients. University hospital Dresden, Germany (DUH) center provide miRNA and mRNA seq data from total plasma extracellular vesicles (EV) of PDAC patients with associated clinical follow information, including 20 benign pancreatic disease patients and 63 pancreatic cancer patients. The four public serum miRNA sequence data containing pancreatic cancer (PC) and healthy control (HC) were GSE106817(2759 PC vs.115 HC), GSE113486(40 PC vs.100HC), GSE112264(50 PC vs.41 HC), GSE109319(24 PC vs. 21 HC). All CT images were acquired using a 64-slice multidetector spiral CT scanner, with a standard slice thickness of 1.0–1.5 mm and a reconstruction interval of 1 mm. All pancreatic cancer patients underwent a standard pancreatic protocol triphasic contrast-enhanced CT examination. For radiomics analysis, images from the portal venous phase were selected due to their consistent clarity in delineating pancreatic tumor boundaries and surrounding vasculature. To ensure data consistency, all imaging data underwent preprocessing, including resampling, intensity normalization of grayscale values, and N4 bias field correction to address potential low-frequency signal inhomogeneities. We use Propensity Score Matching (PSM) method to match the begin and tumor patients from DUH to UMMD & JHC according to age factor. Afterwards, we constructed a new matrix including CT images, EV miRNAs, mRNAs and patients clinical follow up information. The workflow is schematically depictured in Figure 1. Ethical approval to conduct the study for UMMD and DUH after approval by the local Institutional Review Board/ethics committee (UMMD 46/22; 30/01 with amendment 43/14; DUH: EK76032013). Written informed consent from the patients was obtained pre-operatively with the disclosure of research purpose.

**Figure 1:**
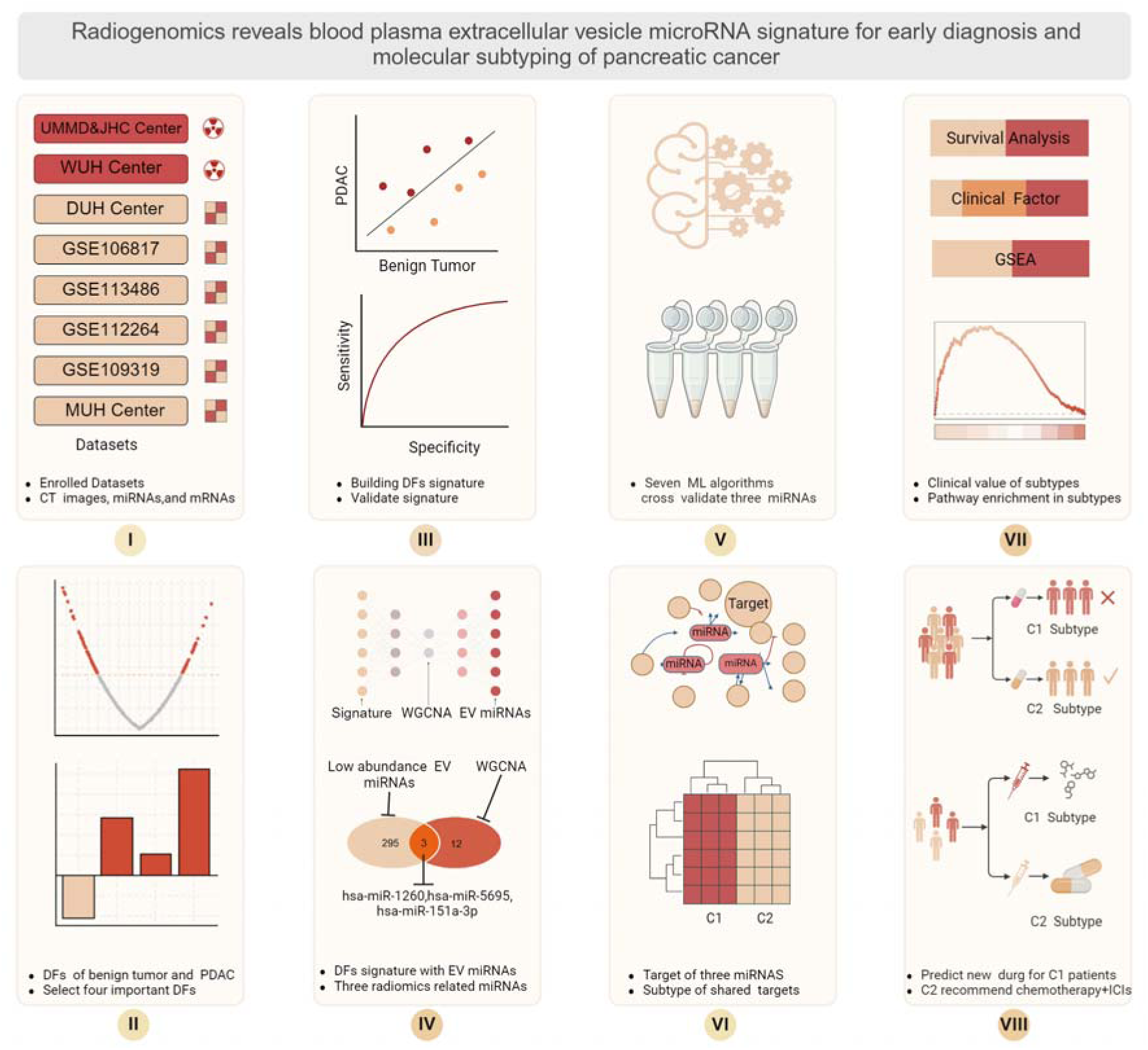
Schematic presentation of the workflow of this study. The use radiomics to aid finding novel EV charged miRNAs to allow PDAC diagnostics.

### EV isolation and RNA sequencing

Details on the protocols for EV isolation including presentation of optical and molecular characteristics of isolated vesicles proofing high quality isolation performance have been described previously by our groups^12^.

EV isolation using ultracentrifugation: 500 μl plasma samples were thawed and mixed with 500 μl PBS. The diluted plasma samples were filtered with 0.2 μm filter and subjected to ultracentrifugation at 100,000×*g*, 2 h, 4 °C in a ultracentrifuge (Sorvall MX150□+□micro-ultracentrifuge, Thermo Scientific, Darmstadt, Germany). The supernatant was removed and the pellet was washed once with ice-cold Phosphate Buffered Saline (PBS, Gibco, Carlsbad, California, USA) and ultracentrifuged again at 100,000×*g*, 2 h, 4 °C. The resulting pellet was resuspended 100 μl PBS and transferred to Vivaspin ® 500 filtration (100,000 MWCO, Sartorius, Göttingen, Germany) for centrifugation at 15,000×*g*, 45 min. The concentrated EVs were stored at −□20 °C until further use for EV characterization.

EV isolation using precipitation method: 550 μl plasma samples were thawed and first centrifuged at 2000×*g* for 20 min. The supernatant was subjected to another round of centrifugation at 10,000×*g* for 20 min. After the second round of centrifugation, 500 μl supernatant was mixed with 250 μl PBS, vortexed and added with 150 μl Exosome Precipitation Reagent. The mixture was incubated at room temperature for 10 min and centrifuged at 10,000×*g* for 5 min. After removing the supernatant completely, the pellet was resuspended with 500 µl PBS and concentrated with Vivaspin ® 500 filtration (100,000 MWCO, Sartorius, Göttingen, Germany) by centrifugation at 15,000×*g*, 45 min. The resulting EVs were stored at −□20 °C until further use for EV characterization.

### RNA sequencing

The eluted EV RNAs were first analyzed for their integrity and concentration using Agilent Fragment Analyzer 5200™ with DNF-472 High Sensitivity RNA Analysis Kit, 15 nt (Agilent Technologies, Santa Clara, California, United States). A range of 1 ng–2 μg RNA was used for complementary DNA (cDNA) synthesis as a preparation for EV RNA sequencing (RNA-seq) libraries with SMARTer smRNA-Seq Kit for Illumina (Takara Bio Inc, Mountain View, California, USA) and were sequenced on an Illumina sequencing platform (NextSeq® 500/550 Mid Output Kit v2, San Diego, California, USA) with run configurations of single read, read 1:51 cycles, index 1:8 cycles, index 2:8 cycles and an average of 3.7 million reads per sample.

Raw reads were first converted from bcl to fastq format using bcl2fastq (v2.20.0.4.422) and subsequently filtered using FastQ Screen to remove potential contaminations by microorganisms or artefacts due to technical issues. The reads were mapped to a phase II reference genome of the 1000 Genomes Project.

### Radiomics Feature Extraction and different features (DFs) analysis

We use 3DSlice soft (www.slicer.org) to mark pancreatic benign and pancreatic cancer in CT images as mask and use original figure as reference. Subsequently, we use python (Version 3.8) soft to extract radiomics feature, respectively. Afterwards, we use limma packages analysis to conduct different feature analysis to identify the significant features between benign and cancer^13^. The p-value ≤ 0.05 was defined as statistical significant.

### Feature selection and radiomics related signature model build

After obtain the DFs, to avoid multicollinearity of the data, we conduct the dimensionality reduction analysis by Boruta algorithms and Lasso regression (LR) model. First, the Boruta algorithms will calculate the importance of each feature and if importance more than shadow feature, it will be selected as important feature, and then, input above important feature into the LR model, the features were selected as significant if the model penalty coefficients were minimized. After selection by machine learning algorithms, we calculate the regression coefficient of each feature in LR model. After that, by combining the feature expression to build the signature model in order to predict the images status from large amount of imaging data. The model formula as listed below, Each regression coefficient of the features is multiplied by its corresponding feature expression level, and then these products are summed. The resulting sum is the Risk Score. The model validation was test with applying WUH center CT dataset, whereas the area under curve (AUC) of Receiver operating characteristic curve (ROC) was used to evaluated the predict ability of model.

### Definition and identification of low abundance EV-derived miRNA transcripts

Low abundance miRNAs are defined as the EV miRNAs with the lowest 30% Counts Per Million (CPM) value across all samples. First, we calculate the CPM value for each miRNA across all samples using the edgeR function^14^. Then, the non-zero miRNAs with CPM values ranked in the lowest 30% of all samples were selected and defined as low abundance miRNAs

### Weighted gene co-expression network analysis (WGCNA) analysis to identify imaging feature related EV miRNAs

According to the radiomics signature, patients will be spilt into low and high-risk group. Low risk classification means images have high percentage of feature parameters from benign images while high risk more likely become cancer featuring images. To explore the correlation between above groups with EV miRNAs, we conduct the WGCNA analysis. The soft-thresholding power of WGCNA was automatically defined by the model, which was then assisting in calculating the expression correlation with a) a given miRNA to obtain gene significance (GS), and b) of module membership with miRNAs to obtain module membership (MM). Based on the cut-off criteria (|MM| > 0.5 and |GS| > 0.1), we obtain the significant miRNAs related to low and high risk images, respectively.

### Hub EV miRNAs identify and validation in serum and tissues level

We merge WGCNA results and low abundance EV miRNAs to identify the candidate hub miRNAs, then we use GSE109319 to validate this EV miRNAs different expression between healthy participants and PADC patients in serum level. In addition, we also validate this EV miRNAs different expression between pancreatic tissue of healthy individuals and PADC tissues in TCGA-sample cohort.

### Screening the best models to aid hub EV miRNAs in blood based diagnosis

We collect serum dataset from GSE106817, GSE113486, GSE112264, our hospital center (UMMD) and GSE109319 dataset to perform this procedure. First, the GSE106817 as training dataset, GSE113486 and GSE112264 are used as independent test dataset. Then we use ten machine learning algorithms (Gradient Boosting Machine GBM ,KNN ,Lasso ,XGBoost ,ENR ,SVM ,LR ,RR ,StepWise ,and QDA) composition to select the best model for prediction. After selection based on highest performance values, we apply our hospital (UMMD) dataset and GSE109319 to validate the model accuracy for ability of clinical prediction.

### Common candidate target mRNAs of hub EV miRNAs

To discover the candidate regulation mechanisms of hub EV miRNAs, we use miRPathDB v2.0 database (https://mpd.bioinf.uni-sb.de/overview.html) to predict the candidate target mRNAs of hub EV miRNAs. Subsequently we select the pass experiment validation mRNAs from the evidence column, as candidate targets of each miRNAs. then, we explore candidate targets of above which were regulated by this three miRNAs at the same time. Finally, we merge above targets and DHU patient’s mRNA sequence to identify the final shared mRNAs which were regulated by three miRNAs at the same time and for the future analysis.

### Cluster of shared target mRNAs and survival analysis between different clusters

We use R package ConsensusClusterPlus to perform the cluster analysis of common target mRNAs, and rank the best cluster results according to the Consensus value output received. Afterwards, we also analyze the survival difference between identified subtypes by R package survival. OS and DFS were used to as event endpoint.

### Clustering of tumor subtype with clinical factors

To explore the clinical value of each subtype, we analyze the relationship between the subtypes and important clinical factors, such as, age, tumor size, number of positive lymph nodes. We also discover the distribution of sex, peri-neural invasion (PNI), and tumor stage the in different subtypes.

### Characterization of immune cell infiltration properties and immune check point activation in each tumor subtype

We use the Microenvironment Cell Populations-counter (MCP-counter) algorithms to calculate immune cell infiltration of each samples and calculated the difference in the two tumor subtypes for each immune cell type separately. We also conduct the relationship between the subtypes and immune check point activation, to predict the candidate subtype could benefit from the immune checkpoint inhibitors.

### Potential drug sensitivity for each subtype

We download drug sensitivity data of molecular characterized cell lines to FDA approved, clinical drugs from GDSC database https://www.cancerrxgene.org/, and then use pRRophetic package to estimate the drug sensitivity of the two discovered subtypes.

### Functional enrichment analysis and pathway prediction

To explore the biological function difference in Biological Process (BP), Cellular Component (CC) and Molecular Function (MF), we conduct the Gene Set Enrichment Analysis (GSEA) analysis. We also use this method to analysis the pathway enrichment difference between the two subtypes. Cluster profile package perform this operation and set p-value ≤ 0.05 as significant enrichment results.

## Results

### Enrolled population and baseline information for CT images and EV miRNA

A total of 272 patients enrolled this study providing CT imaging, including 173 in UMMD&JHC center while 99 in WUH center. In UMMD&JHC center, also including 46 pancreatic benign lesion and 127 pancreatic cancer CT images. Most of pancreatic patients in this center are older with obesity, but less smoking or alcohol and less with diabetes. For WUH center, most pancreatic cancer are female, and also with a high account for older, and obesity. The DUH provide the patients with EV-miRNAs, EV-mRNAs data and follow up information. About 82.5% (52/63) patients are older, and 34 patients are female. The clinical characteristics of the data cohorts are summarized in Figure 2.

**Figure 2:**
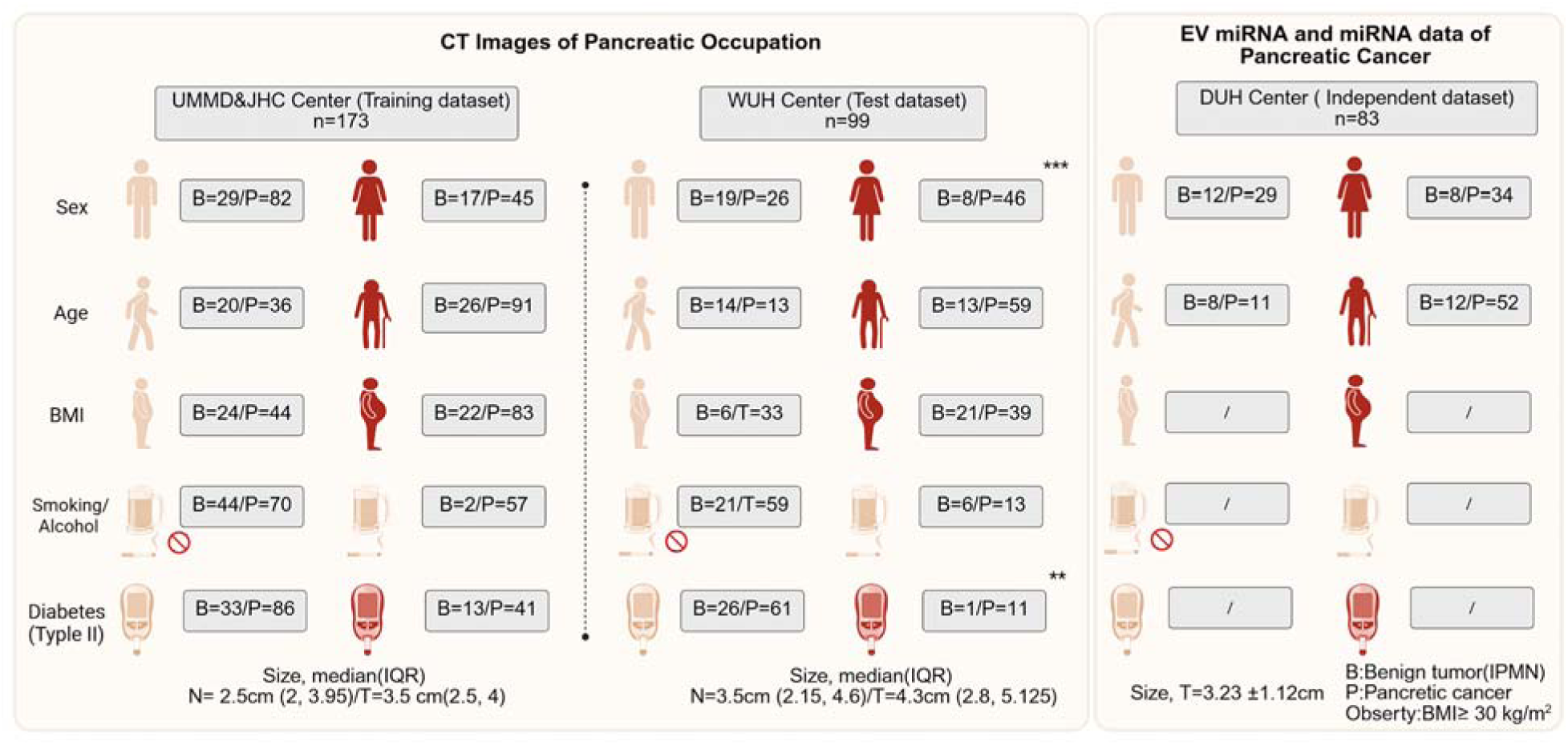
The base line information of clinical parameters of patients enrolled from four centers in this multi-center trial.The * means p less than 0.05,** means p less than0.01,and ***means p less than0.001.

### Different expression radiomic features between pancreatic benign lesions and aggressive tumors

Before the analysis, we conduct the PSM procedure to match the benign and aggressive tumor from DUH to JHC, respectively, according to age. After match, we found both center baseline difference are removed (Figure 3A). Then the difference expression radiomics features was conduct, the results indicate that a total of n=88 significant features demonstrate differences between groups (Figure 3B).

**Figure 3:**
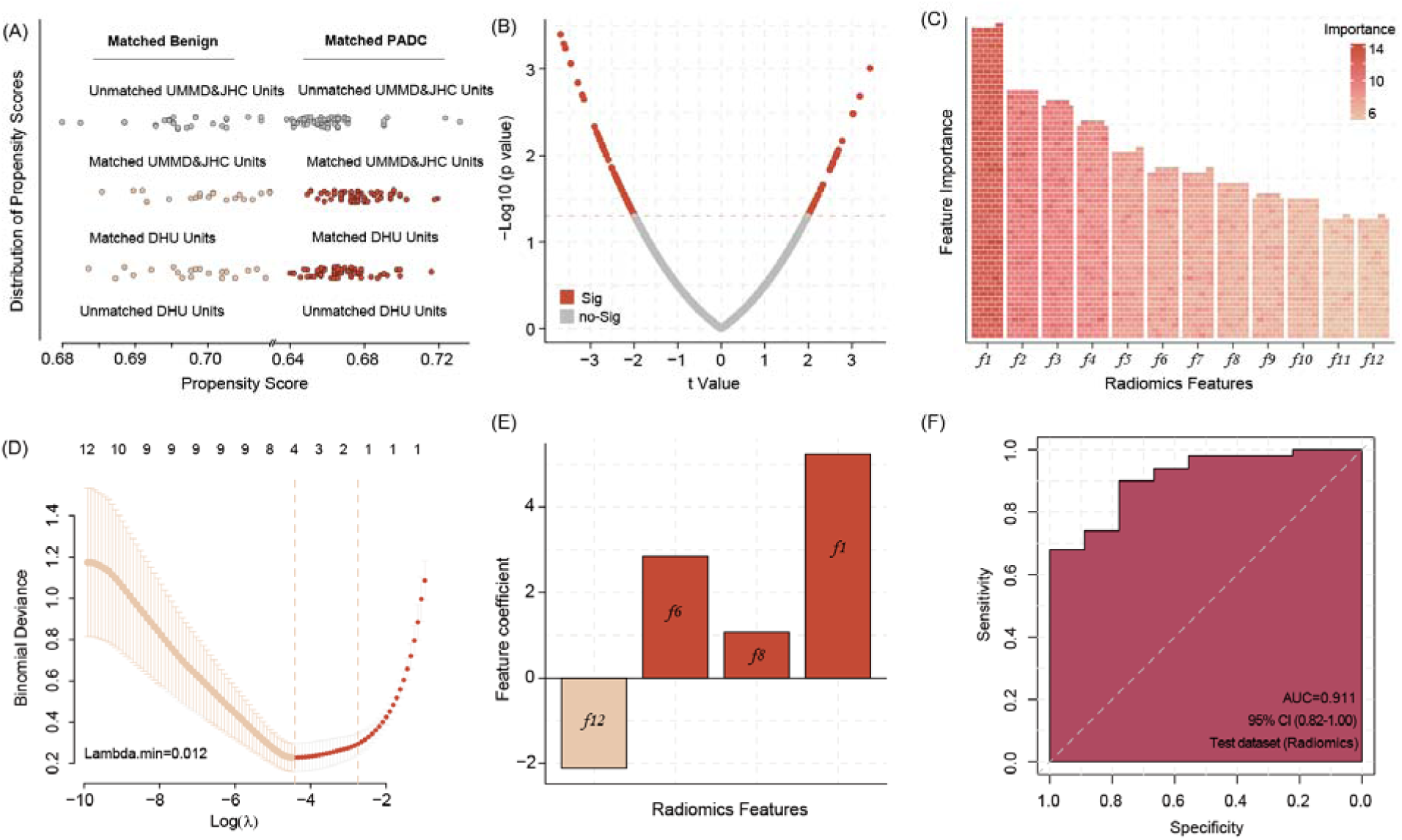
Propensity Score Matching (PSM) allows matching data of benign pancreas lesions and PDAC patients from DUH to UMMD & JHC patients according to the age factor, all of DUH patients successful matched similar patients (A). The different radiomic features between the benign lesions and PDAC patients CT images(B). 12 most important radiomic features differentiating between the benign pancreatic lesion and PDAC patientś CT images identified by the Boruta algorithms (C) Four radiomic features were selected by Lasso Regression to build model signature (D&E). Applying the four radiomic features related signature in image analysis show high accuracy in predicting the PDAC manifestation in the WUH test dataset(F).

### Four important radiomic feature was selected to build a related signature

We use Boruta algorithms to select the important features and result show that a total 12 features were identified(Supplement Table 1), which more than shadow features. In addition, after input above features into LR algorithms and we can found that four features are list as key features(Figure 3C-D). Based on LR model with regression coefficient and feature expression, we build a four radiomics feature related signature and validate the prediction ability in WUH center data, revealing a signature accuracy of prediction efficiency (AUC=0.911) (Figure 3E-F).

**Supplement Table 1.**
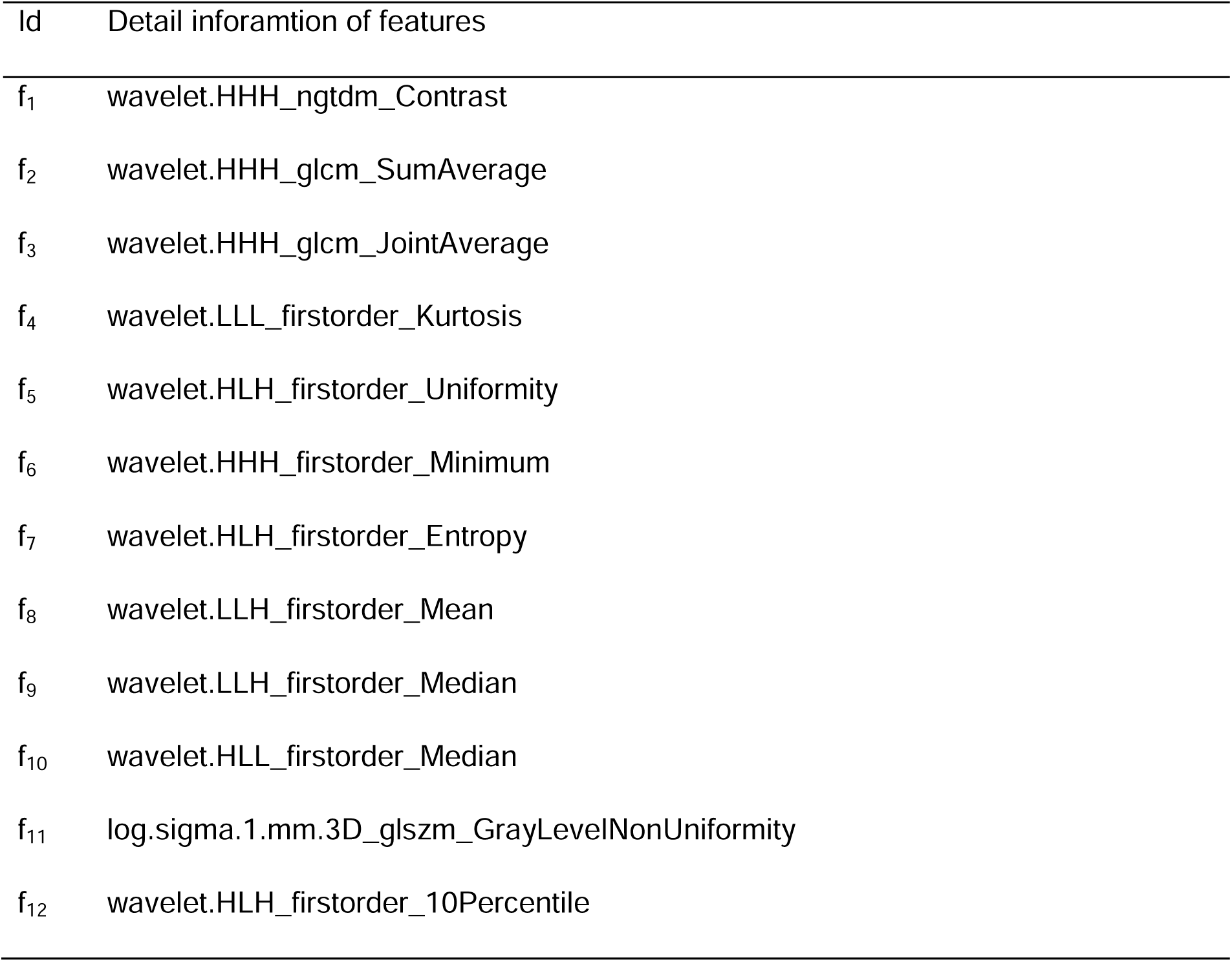
Important radiomic features.

### Three EV miRNAs are associated with radiomic features

After radiomics signature was build, each patients presents an individual risk score and we split patients into high-risk and low-risk patients, according to median value of risk score. We use WGCNA to connect the EV miRNA data and two imaging-featured patient risk groups. According to the WGCNA analysis, the green module was identified as the key module and includes 12 hub co-expressed miRNAs. The number of low abundance of miRNA are calculated was n=295. Merging both results, three miRNAs are identified (hsa-miR-1260b, hsa-miR-151a-3p and hsa-miR-5695) and selected for subsequent alignment with associated with radiomic features (Figure 4A-C).

**Figure 4:**
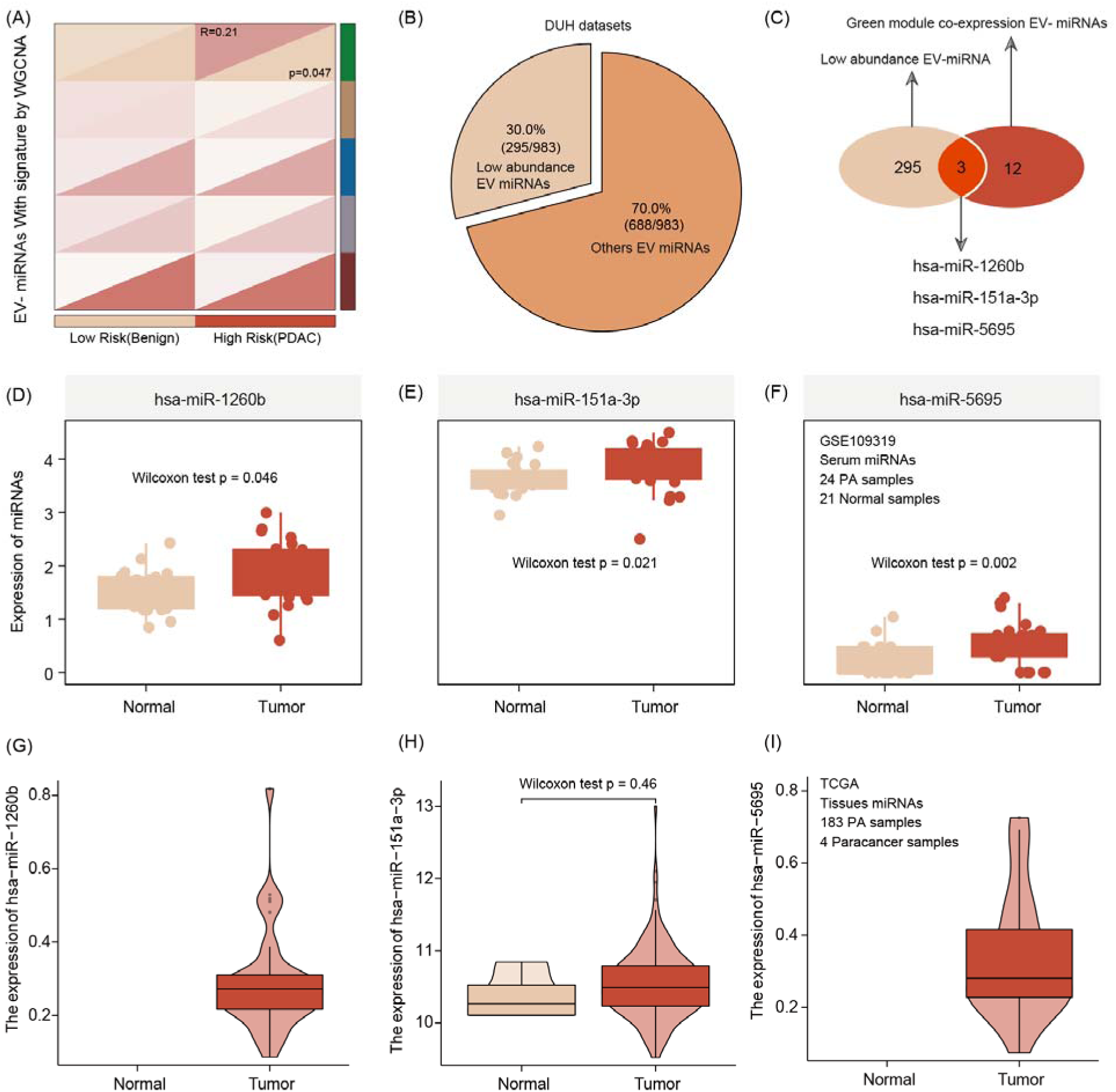
EV miRNA presenting the risk group stratification based on radiomics signature by WGCNA analysis featuring green mode discovering our key module for further analysis (r=0.21, p=0.047) (A). The number of low abundance miRNA in the entire EVseq dataset cohort is n= 295 (B). Out of those low abundance miRNAs, n=12 present matching candidates differentially expressed in high risk group patients. Alignment to our radiomics feature parameters identified three core miRNAs (hsa-miR-1260b, hsa-miR-151a-3p and hsa-miR-5695) (C). The three key miRNAs show significantly different expression levels in tumor condition, both for serum (D-F) and tissue (G-I).

### Expression validation of three EV miRNAs

We use serum and tissues sample to validate the expression of three hub EV miRNAs and the results show that compare with healthy serum sample, this three miRNAs are enriched in tumor patient serum sample. Interestingly, this correlation of upregulation in tumor conditions was also true when comparing tumor tissue with non-tumor tissue (Figure 4D-I).

### Three EV miRNAs predictd pancreatic cancer with high accuracy

We use seven machine-learning combos to train and test the ability of three EV miRNAs levels to predict tumor manifestation and clinical course of patient. The results show that in the GBM-default (cutoff=0.75) model, three EV miRNAs show a high accuracy to predict the pancreatic cancer with the training accuracy was 0.978 with AUC=0.978, and two test dataset accuracy are 0.923 with AUC=0.919, and 0.871 with AUC=0.857,respectively. Then, we choose GBM model for the extend validation by our hospital data(MUH) and GSE109319.Before this procedure, we use combat package to remove the batch effect allowing the merge of the two datasets. The results of the extend dataset validation via GBM further highlights the high diagnostic accuracy of the three EV miRNAs (accuracy =0.894, and AUC=0.897) (Figure 5), while the CA19-9 predict auc is 0.843(95% CI, 0.762−0.924).

**Figure 5:**
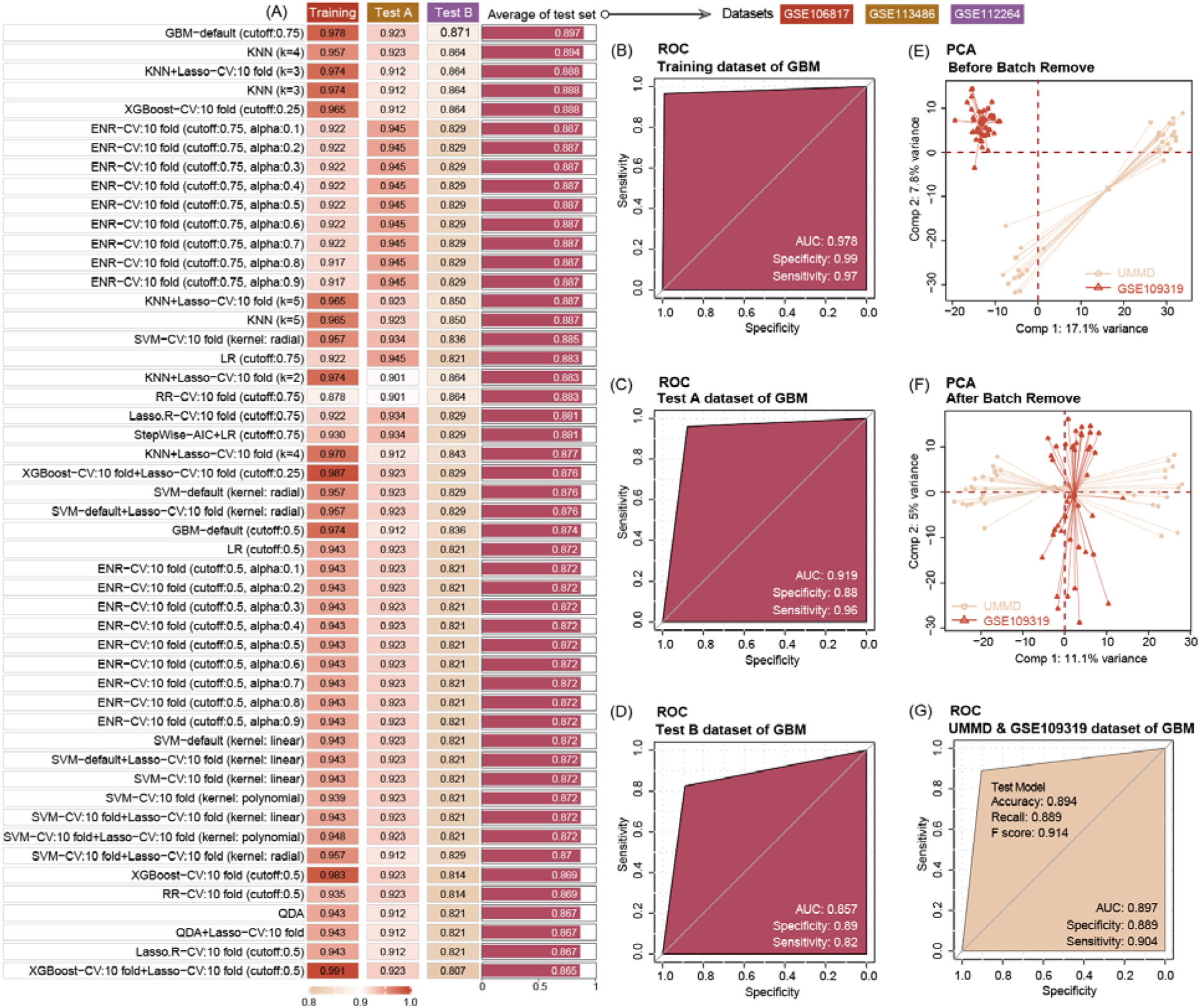
Ten machine learning algorithms demonstrate that three key miRNAs show a high accuracy to diagnosis PDAC in early stage, no matter in training or test datasets. The best machine learning algorithms is GBM (cutoff:0.75) (A). Three miRNAs prediction ability of training dataset (GSE10106817) in GBM model is 0.978(B). Three miRNAs prediction ability of test dataset (GSE113486 and GSE112264) in GBM model is 0.919, and 0.857, respectively(C&D). Data distribution before removal of batch effect of our center data and GSE109319 dataset (E). Data distribution after remove batch effect of our center and GSE109319 dataset (F). Three EV miRNAs prediction ability to identify cancer of our center data and GSE109319 in GBM model is 0.897(G).

### Identification of two molecular subtypes of PDAC with significant clinical predictive value

After predicting molecular targets of three miRNAs, we obtain a number of 117 mRNA are shared candidate targets (**Supplement Table 2**). Stratifying the tumors according to the level of expression of those allows the differentiation of tumors in cluster into two subtypes termed Cluster 1 (C1) and C2 (Figure 6A and Supplement Figure 1). The survival analysis show that compared with C1, the C2 patients feature a prolonged survival outcome, no matter in OS or DFS(Figure 6 B&C). In addition, analysis the relationship with clinical factors of patients, we also found the C1 subtype patients are older in age and carry bigger tumor sizes and higher number of tumor cell positive lymph nodes(Figure 6D-F), while compared with C2 subtype patients. We also demonstrate C1 patients particular aggressive in female patients, feature high number of tumors containing perineural invasion (PNI) and advanced pathological tumor staging (Figure 6 G-I).

**Figure 6:**
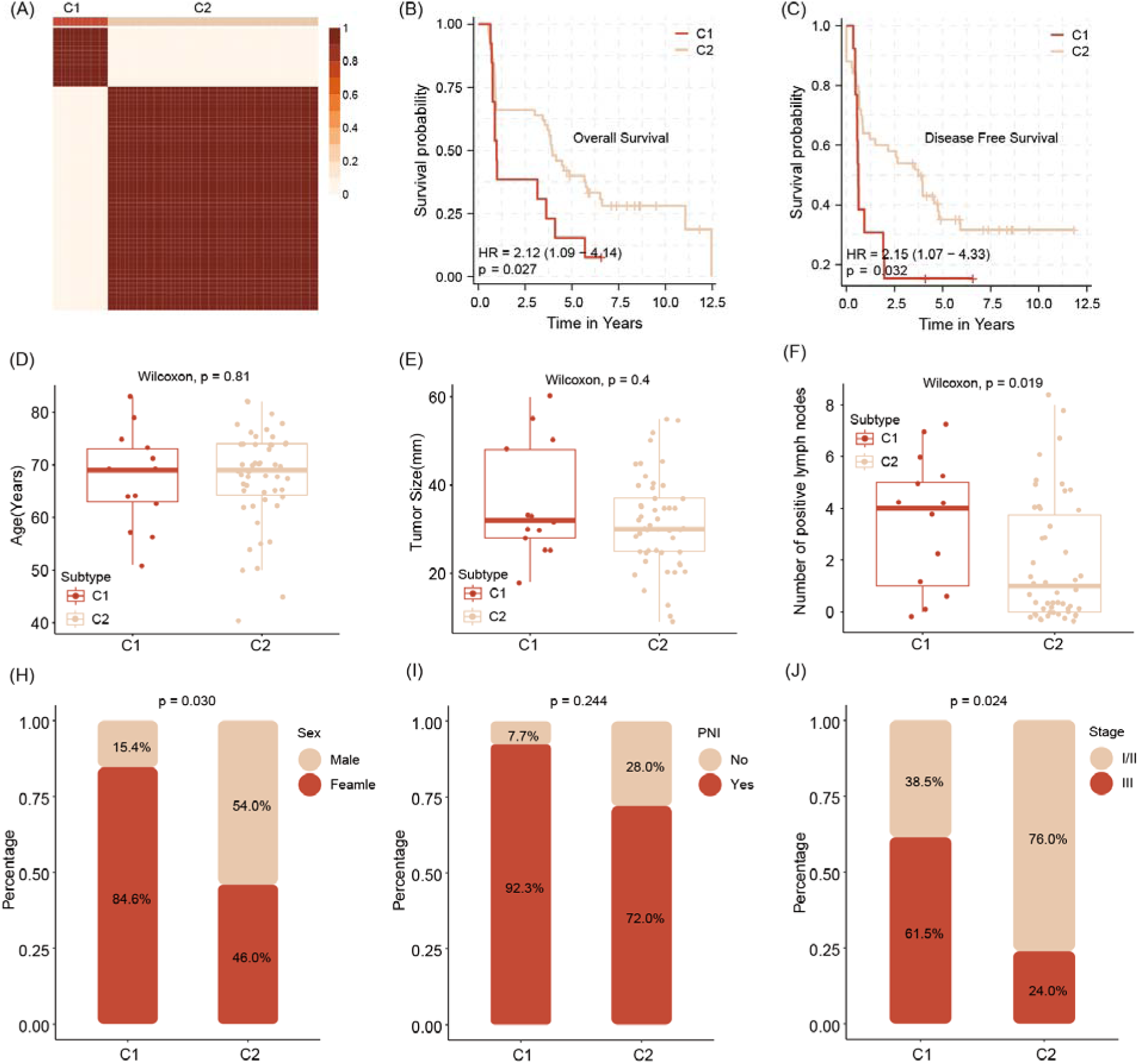
Stratification of abundancy levels of shared mRNAs by Non-negative matrix factorization method allows the clustering of patients into two subtypes (C1 and C2) (A). Patients of the C1 with a poor out outcome in OS and DFS(B&C). C1 subtype patients are characterized with older age and bigger average tumor size, higher number of tumor cell positive lymph nodes as compared to C2 patients (D-F). C1 patients are predominantly female, have tumors with pathological classification marks of perineural invasion and advanced tumor stage (G-I).

**Supplement Table 2.**
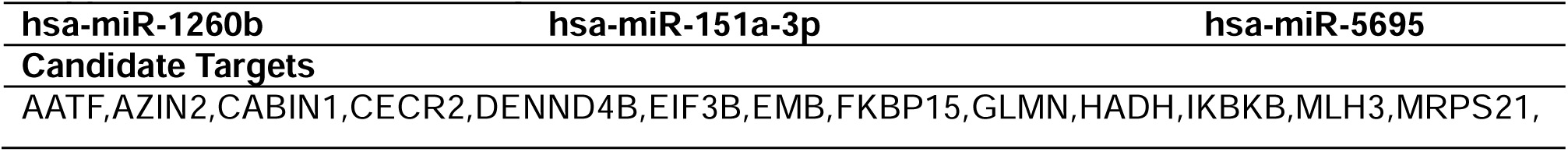
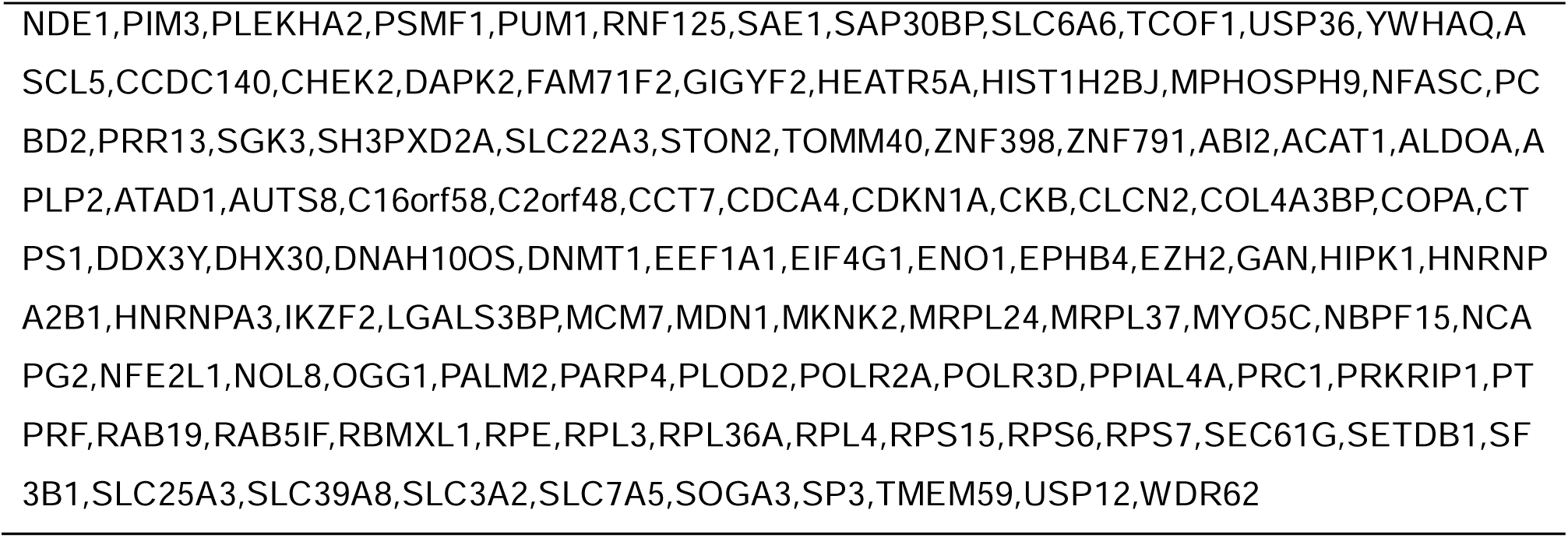
Shared targets of three EV miRNAs.

### C2 patients with high immune infiltration and positive with immune checkpoints

Testing for gene signatures that indicate immune cell existence, we show that C2 tumors are significantly more enriched for immune cell signals, including those for CD8 T cells, Cytotoxic lymphocytes and NK cells (Figure 7 A&B). In addition, samples of patients of that subtype show also a more physiological expression signature in terms of immune checkpoints as compared to C1 tumors (Figure 7C).

**Figure 7:**
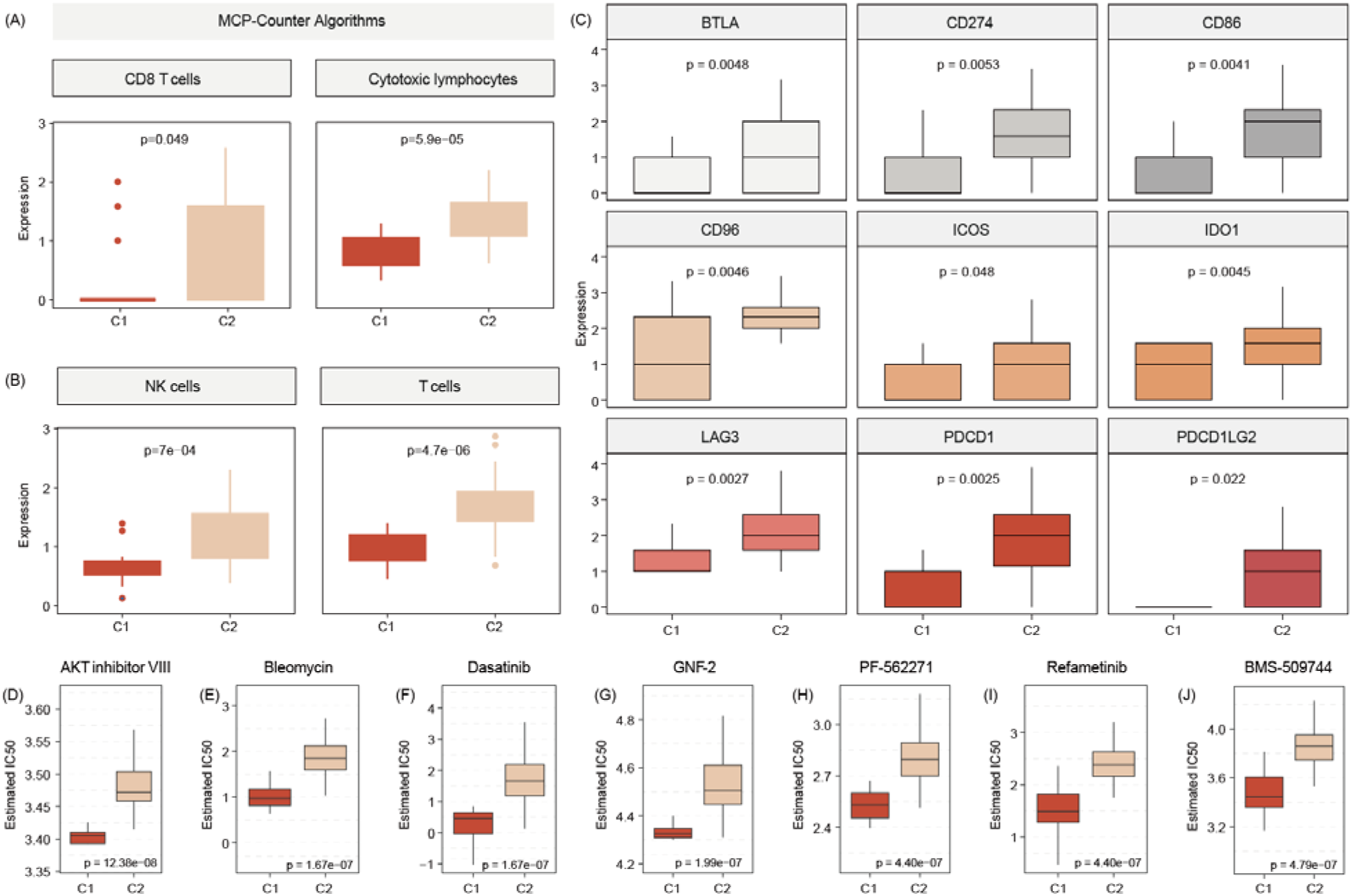
C2 subtype is positively associated with elevated levels of transcripts regulating gene pathways encoding for CD8 T cells, cytotoxic lymphocytes, NK cells(A&B). Commonlyknown immune checkpoints also higher expressed in C2 subtypes(D). Aligning in vitro drug sensitvtiy and expression data from the GDSC database to subtype signature, predicts that tumors of C1 patients might be more sensitive to AKT INHIBITOR VIII, Bleomycin, Dasatinib, GNF-2, PF - 562271, Refametinib, BMS-509744 as C2 subtypes.

### Prediction of drug sensitivity of C1 patients

We use Genomics of Drug Sensitivity in Cancer (GDSC) data to merge cell line response data and expression data with expression signals in C1 and C2 tumors. This analysis indicated that C1 patients may benefit from therapy with FDA approved AKT inhibitor VIII, Bleomycin, Dasatinib, GNF-2, PF −562271, Refametinib, BMS-509744 (Figure 7E).

### Functional annotation and pathway enrichment for C1 subtype

GO functional enrichment analysis indicated that the C1 subtype is enriched for intermediate filament organization, GOCC ribosome, as well as GOMF symporter activity (Figure 8 A&B). Pathway enrichment analysis showed that the C1 subtype was activated with the Reactome fatty acids, Reactome diseases of metabolism, as well as Reactome biological oxidations pathways, and may be inhibited with Reactome apoptosis, Reactome DNA repair, and Reactome signaling by Hippo pathways (Figure 8C).

**Figure 8:**
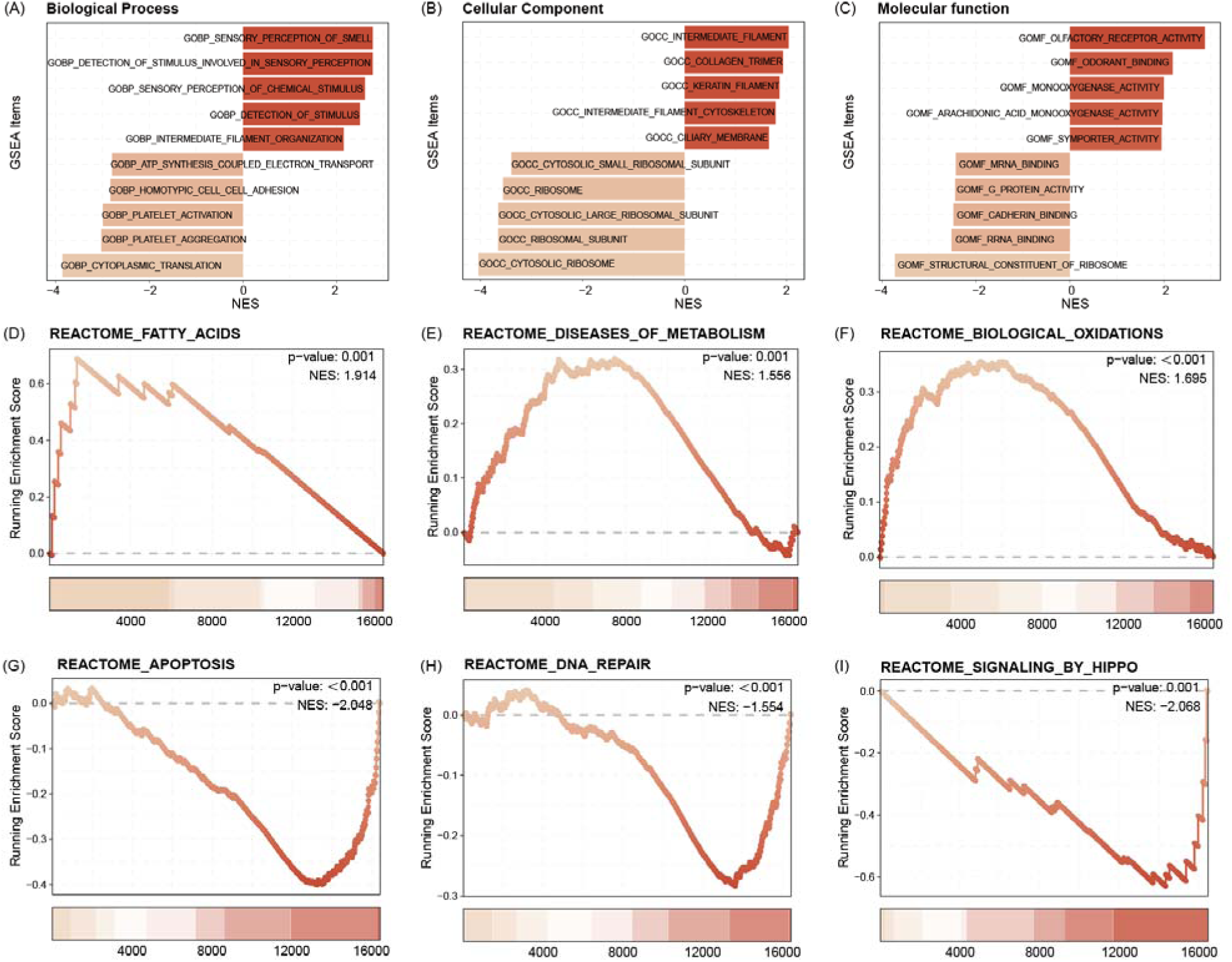
GO functional enrichment analysis indicated that the C1 subtype is enriched for intermediate filament organization, CC ribosome, as well as MF symporter activity(A-C). Pathway enrichment analysis showed that the C1 subtype was activated with the Reactome fatty acids, Reactome diseases of metabolism, as well as Reactome biological oxidations pathways, and may be inhibited with Reactome apoptosis, Reactome DNA repair, and Reactome signaling by Hippo pathways (D-I).

## Discussion

Using multicenter radiomics of Asian and Western world patients, we identified 3 plasma total extracellular vesicle fraction microRNAs that demonstrated strong predictive performance and prognostic value, offering new insights for noninvasive early diagnosis and prognosis of pancreatic cancer. This study is the first to distinguish between benign lesions and pancreatic cancer by identifying novel and significant plasma extracellular vesicle microRNAs based on differential radiomics features. We focused on low-abundance microRNAs, which are often overlooked in routine RNA sequencing analyses. These 3 microRNA candidates were validated across multiple cohorts, achieving excellent predictive performance in both testing and external validation sets. Additionally, these microRNAs demonstrated prognostic value, potentially aiding in treatment selection.

Pancreatic cancer remains one of the most lethal diseases, with insufficient early detection methods and limited treatment options^15,16^. This study aimed to develop a liquid biopsy nucelaic acid diagnsotic test based in EV-derived microRNA using radiogenomic analysis to enhance diagnostic accuracy and molecular subtyping of pancreatic ductal adenocarcinoma. By focusing on imaging features to distinguish benign lesions from pancreatic cancer, we identified underlying genetic factors contributing to these differences. Unlike previous studies on serum microRNA^17,18^, our research identified three microRNAs through radiomics that reflect tumor-specific changes rather than systemic variations. Additionally, our findings reveal relationships between imaging features and tumor biology, improving diagnostic accuracy and offering a more comprehensive tool for early detection and personalized disease management.

Although numerous studies have reported on microRNA sequencing results^19^, highly abundant microRNAs often mask the signals of many others that are present at very low abundance during routine analysis. Furthermore, if a microRNA is associated with tumor development, it is likely to be in low abundance during early detection and may even be undetectable in benign lesions. In our study, after identifying 12 microRNAs related to radiomic differences, we further defined three (hsa-miR-1260b, hsa-miR-151a-3p and hsa-miR-5695) of them as being of low abundance. These microRNAs showed significantly higher expression in pancreatic cancer blood than in non malignant cancer patients’ plasma, with hsa-miR-1260b and hsa-miR-5695 undetectable in pancreatic cancer samples. The model based on these 3 microRNAs demonstrated strong performance in the training group (AUC, 0.978; sensitivity, 0.97; specificity, 0.99), testing group (AUC, 0.919; sensitivity, 0.96; specificity, 0.88), and external testing group (AUC, 0.897; sensitivity, 0.90; specificity, 0.89).

Previous studies on microRNAs as diagnostic biomarkers for pancreatic cancer vs healthy controls report AUCs of 0.78-0.96 and sensitivities of 0.62-0.88^19^, but many lack external validation^20,21^. One case-control study identified 2 diagnostic panels based on microRNA expression (index I: 4 microRNAs; AUC, 0.86; sensitivity, 0.85; specificity, 0.64; index II: 10 microRNAs; AUC, 0.93; sensitivity, 0.85; specificity, 0.85) ^22^. Our study achieved similar performance using only 3 microRNAs, making it more practical for clinical application.

To the best of our knowledge, hitherto none of the three identified microRNAs have been reported in the context of diagnostic biomarkers in pancreatic cancer. Moreover, to our knowledge, hsa-miR-5695 only indirectly been linked to breast carcinogenesis^23^ and subtypes of metastatic prostate cancer^24^, while hsa-miR-1260b has been implicated in promoting tumorigenesis in lung cancer^25^, breast cancer^26^, sarcoma^27^ and prostate cancer^28^. Interestingly, in a screen of microRNAs that are differentially expressing in total plasma fraction of patients suffering from intraductal papillary mucinous neoplasm (IPMN) as compared to non-diseased controls, hsa-miR-1260b was identified as one of the top upregulated signals in plasma of IPMN cases^29^. Our work further enforces the potential of hsa-miR-1260b abundancy in plasma to detect onset of abnormal pancreas genesis. Exosomal hsa-miR-151a-3p has emerged as a potential novel biomarker for predicting bone metastasis in lung cancer^30^. A very recent report of results of multicenter trial in Asian participants also identified circulating 2’-O-methylated form of hsa-miR-151a-3p is upregulated in total fraction of plasma of PDAC patients compared to those derived from healthy donors or patients with chronic pancreatitis^31^. Focusing on EV fraction, our results verifies the potential of this newly discovered biomarker in pancreatic cancer blood diagnosis.

Pancreatic cancer currently has only two standard systemic treatments with limited efficacy, and the patient population responsive to immunotherapy and targeted therapy remains unclear^32^. Therefore, we explored the prognostic value of these microRNAs, which allowed for patient stratification. Patients classified as C2 had better prognoses, with higher immune cell infiltration and more immune checkpoint expression, suggesting they may be suitable for immunotherapy after standard treatment. In contrast, patients classified as C1 had poorer prognoses, related to inhibited pathways such as DNA repair and apoptosis.

The findings suggest potential for developing kits for early detection and treatment stratification, and highlight these three candidates for further exploration. However, despite thorough matching of imaging, genomic, and clinical data, biases may still exist. Additionally, using the MCP-counter algorithm for blood samples is a limitation of our study, as it was originally designed for solid tissues, which may affect the accuracy of cell population quantification in the blood. Furthermore, when validating the three microRNAs in tissue, the sample size of paracancerous tissue was limited. Lastly, since our EV purification did not use measures to enrich for cancer cell derived EVs such as CD147 membrane presenting amplitude^33^, we cannot exclude contamination of our miRNA data due to signals from non-cancer cell background, which might impact the cancer specificity of our proposed blood test. Moreover, our data is based on OMICS data acquisition. Real world application of our proposed test would benefit from proofing the predictability and sensitivity of the miRNAs by targeted methods, such as RT-qPCR or CRISPR/Cas diagnostics and in prospective, multicenter trial. In addition, We acknowledge the absence of third-phase validation results, which will limit application in the clinical practice.

## Data availability

All data generated from this study, if not included in this article, are available from the corresponding authors on reasonable request.

## Funding

The MD fellowship program of the medical faculty of University Magdeburg supported the project.

## Authors contribution

Data acquisition and experiments, data analysis, layout: WS, IZ, GZ,

Resources: UDK, CK; RSC, GR, MP, GLZ

Data interpretation, writing and review manuscript: all authors

---

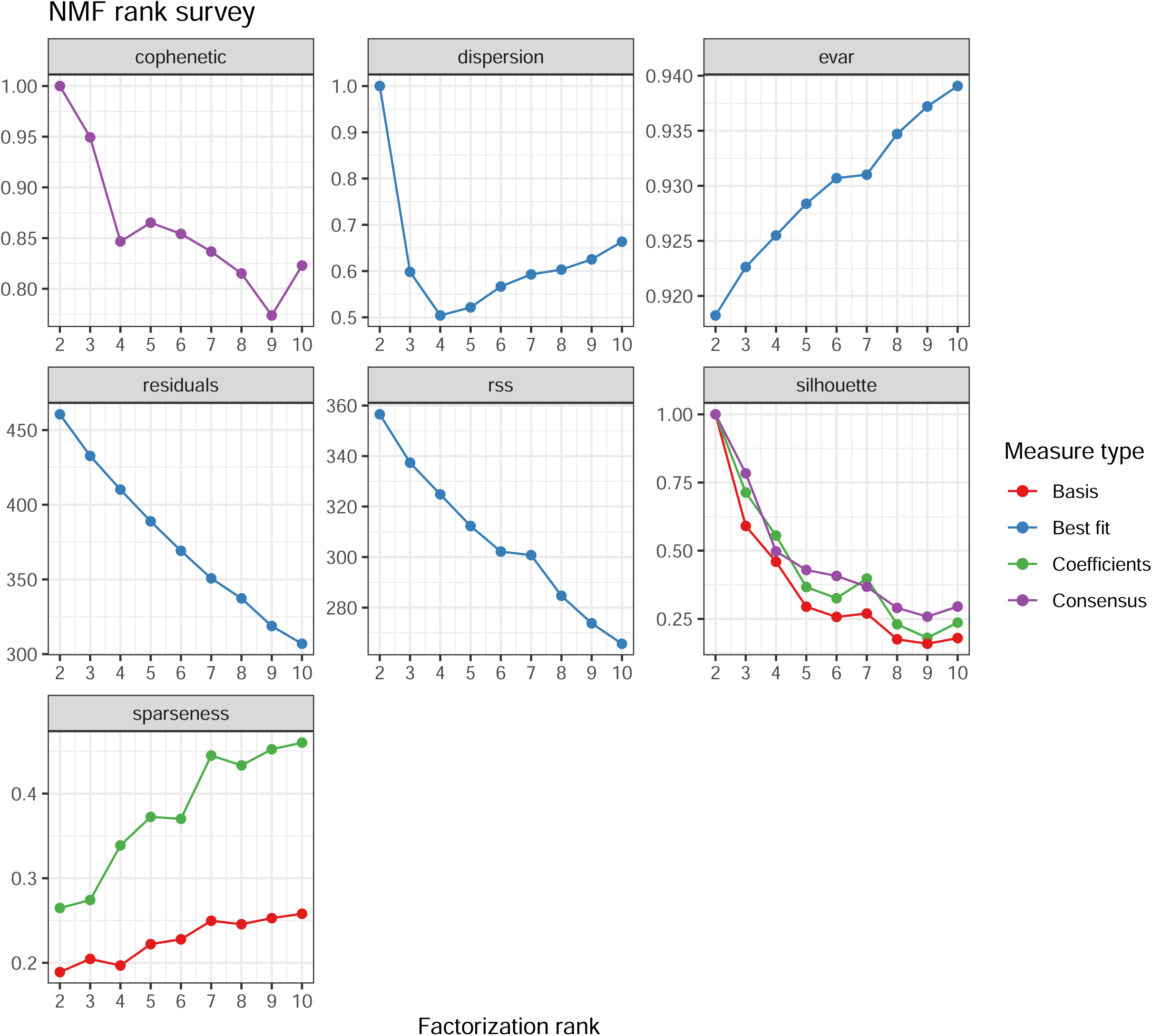

## Notes

The authors declare no potential conflicts of interest

### Competing Interest Statement

The authors have declared no competing interest.

### Funding Statement

This study did not receive any funding. The work of JN was supported by the scholarship for doctoral project from the medical faculty Magdeburg, Germany.

### Author Declarations

The data from plasma samples and associated clinical parameters of enrolled patients were provided by university hospital Dresden (DUH), comprising collection of three cohorts from German centers. Plasma samples from the enrolled patients were received from the (1) Department of Visceral, Thoracic and Vascular Surgery, University Hospital Carl Gustav Carus Dresden, Dresden, Germany; (2) Department of General, Visceral and Transplantation Surgery, University Hospital Heidelberg, Heidelberg, Germany; and Department of Surgery, University Hospital Erlangen, Erlangen, Germany after approval by the local Institutional Review Board/ethics committee (Dresden: EK76032013; Heidelberg: 159/2002; Erlangen: 156_19B). Written informed consent from the patients was obtained pre-operatively with the disclosure of research purpose. CT imaging data and associated clinical parameters of patients was retrieved from university hospital Magdeburg, Germany (studies #33/01 with current amendments from year 2024, and #46/22 approved by ethical committee of University Magdeburg), The Affiliated Hospital of Jiaxing University, China (study # 24-LY-674 approved by ethical committee of the The Affiliated Hosptial of Jiaxing University) and Wannan medical university hospital, China (for retrospective studies not involving human samples, local rules in Anhui province do not require an dedicated ethical approval to conduct the study, an exemption letter was provided).

### Summary of Updates

We provide an in depth revision of the manuscript addressing the reviewer critics we received.

